# ExonViz: A user friendly application to visualize transcripts and genetic variants

**DOI:** 10.1101/2024.09.18.24313945

**Authors:** Redmar R. van den Berg, Marlen C. Lauffer, Jeroen F.J. Laros

## Abstract

Visualization of genes and genetic variants as well as transcript structure is essential within the human genetics community. Such illustrations represent a key tool in communicating genetic concepts and facilitating discussions on therapeutic interventions.

There currently are no easily usable tools which allows the users to draw all features required for a comprehensive overview of a transcript’s structure and the localisation of variants of interest.

Here we introduce ExonViz, an online application that creates biologically accurate transcript figures, including features such as coding regions, genetic variants and exon reading frames. Transcript definitions are automatically retrieved from Ensembl and RefSeq. We illustrate the full functionality of ExonViz by generating a figure for all variants reported in ClinVar for *CYLD*. ExonViz is available online via the Dutch Center for RNA Therapeutics website and can be installed locally via PyPI.

## Introduction

Visualization of transcripts, including features like coding and non coding regions, reading frames and the mutational landscape is important within the field of clinical and human genetics (Walker et al., 2023). Illustrating the exon structure and the distribution of variants over the gene is common practice, especially when new genes or transcripts have been discovered. These illustrations are also used to assess potential genetic treatment options (e.g., canonical exon skipping), in teaching settings, in diagnostics, to identify mutational hotspots and for genetic counseling.

To date, most people have to resort to manually drawing transcripts with tools like Illustrator, Photoshop or BioRender, or forgo illustrations altogether. Some tools have been made available that aid in drawing transcripts (ggtranscript (Gustavsson et al., 2022) and wiggleplotr (Alasoo, 2017)), visualize different transcript isoforms (genepainter (Mühlhausen et al., 2015)), or visualize variants (Variant View (Ferstay et al., 2013)). However, none of these tools can automatically draw exon reading frames.

Knowledge about the exon frames aids in the assessment of the pathogenicity of genetic variants using the ACMG-AMP guidelines (Richards et al., 2015) when evaluating exon spanning deletions (Cheerie et al., 2025) and when interpreting the effects of splice altering variants (Walker et al., 2023). Creating transcript visualizations must be quick and easy if they are to be utilized in clinical and day to day settings, rather than to create a bespoke figure for a manuscript or presentation.

To this end we present ExonViz, an application that automatically creates biologically accurate transcript visualizations, requiring minimal user input. By using arrows, notches and straight edges to indicate the reading frames of exons, it helps users identify which exons share common reading frames. This aids in genetic counseling, genetic therapy assessment and in educational settings.

## Availability

To facilitate ease of use, ExonViz is available as a web application on the website of the Dutch Center for RNA Therapeutics^1^. The web interface features the most commonly used options with sensible default values. For more advanced uses, ExonViz is also available as a Python programming library accompanied by a command line interface, which can be installed via PyPI^2^. Extensive documentation on ExonViz is available on Read the Docs^3^. Figures generated by ExonViz fall under the Creative Commons BY license^4^, which means they can be shared and adapted as long as appropriate credit is given. The source code itself is available on Github^5^ under the AGLP-3.0 license.

The command line interface allows for more fine grained control over both the visualization parameters (such as exon and variant colors and names) as well as the transcript structure, allowing users to modify existing transcripts or to specify their own. This is particularly useful when illustrating the effect of splice altering variants, novel transcripts and the distribution of variants across a transcript, as can be seen in the online documentation^6^.

## Usage

In Figure 1A, an overview of a typical ExonViz visualisation is shown, with the important features highlighted. ExonViz will render the coding regions and indicate variants using a colored pin, using different colors to distinguish the variants.

**Figure 1:**
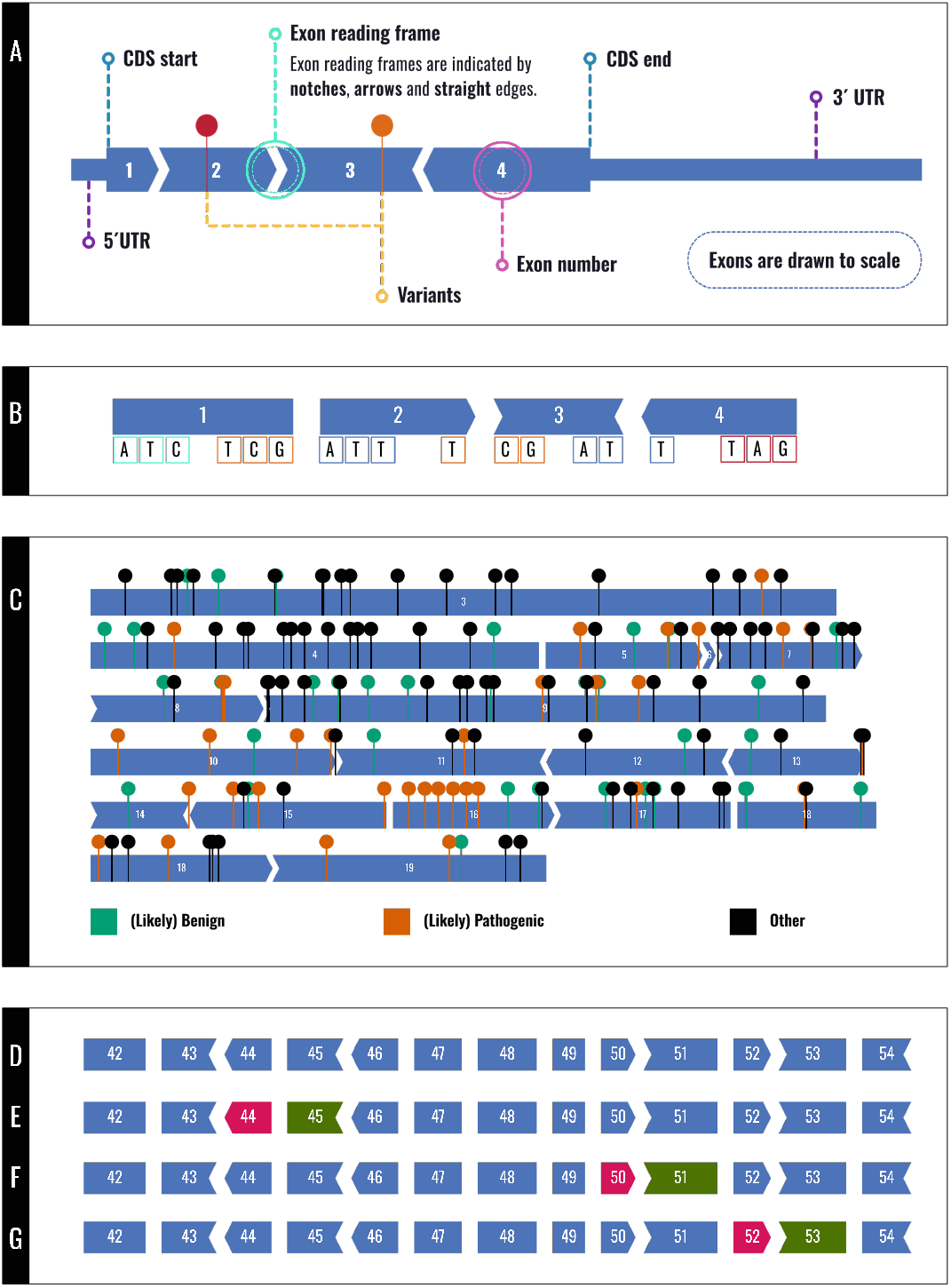
**A** Overview of the features included in an ExonViz figure, based on *SDHD* (NM_003002.4); **B** Illustration of the exon reading frames, with codons indicated in alternating colors; **C** All ClinVar listed variants the coding region of *CYLD* (NM_001378743.1). Likely benign and benign variants are depicted in Bluish green, likely pathogenic and pathogenic variants are depicted in Vermilion and any other variant is shown in black; **D-G** Partial coding region of transcript Dp427m of the *DMD* gene (ENST00000357033.9). Deleted exons are indicated in magenta, skipped exons are indicated in dark green; **E** Deletion of exon 44, which can be rescued by Casimersen skipping exon 45; **F** Deletion of exon 50, which can be rescued by Eteplirsen skipping exon 51; **G** Deletion of exon 52, which can be rescued by Viltolarsen skipping exon 53.

The default settings can be modified to customize the figure, for example by hiding the non-coding region as is shown in Figure 1B. It is also possible to modify thee shape of the variants to a less intrusive `bar` instead of the default `pin`, and the `color` of the exons can also be configured. Finally, the `height` of the exons can be set, which also influences the size of the arrows and notches. Since the figure has been scaled to fit on the page, increasing the `height` gives the transcript a more stocky appearance.

The only required input is a gene name, transcript identifier or HGVS variant description. If a gene name is specified, ExonViz will automatically determine the corresponding MANE Select transcript (Morales et al., 2022), since this is usually the most relevant transcript (Pozo et al., 2022). In cases where multiple MANE Select transcripts have been defined for a gene, the first one will be selected. ExonViz will automatically fetch annotations like exon structure and coding region for the selected transcript. By default, only the coding region of the transcript will be shown, but rendering of the non coding regions can be enabled. The non coding regions will be drawn at half the `height` of the coding region. If any variants were specified, they will be indicated in the correct location, as can be seen from Figure 1A and 1C.

The transcript layout can be modified using the `scale` and `width` parameters. The `scale` parameter determines how many pixels should be used per base pair (default is 1), while the `width` parameter determines the width of the page. Exons that are too large to fit on the page will be split over multiple rows. Exons can also be split if they are close to the end of the page (e.g., exon 18 in Figure 1C). Since the arrows and notches take up a certain amount of space, small exons have to be drawn at a scale larger than 1, depending on the `height`. If this is the case, the user will be notified of the minimum scale at which the transcript can be drawn.

The variants specified in the HGVS description will be visualized at their correct location, as long as the *first* position of each variant is located inside an exon. Only variants that start inside an exon will be included in the figure, otherwise the variant is discarded and a warning will be shown to the user. This means that a big deletion that starts in an exon and includes part of an intron will be shown, whereas a big deletion that starts in an intron and includes part of an exon will be removed. When any variants are drawn on a transcript, a legend is added at the bottom of the figure to indicate the color and name for each variant.

## Materials and Methods

ExonViz is written in Python 3, its web interface is built using Flask. It uses the public Mutalyzer API (Lefter et al., 2021) to fetch transcript information. This gives ExonViz access to all transcripts defined in the RefSeq (O’Leary et al., 2016) and Ensembl (Harrison et al., 2023) databases across many species, ranging from human and mouse to fruit fly and coelacanth. Transcripts and annotations defined on the reverse strand are reversed on the fly, so ExonViz always visualizes transcripts in their forward orientation.

The start and end frames, which refers to the alignment between the exon boundary and the codon boundaries, are indicated by the shape of the exon, as can be seen in Figure 1B. If the first base of an exon is also the first base of a codon, the start frame of the exon is 0. If an exon starts at the second base of a codon, the start frame is 1, etc. The same holds for the end frames.

As shown in Figure 1B, exon boundaries in frame 0 are drawn with a straight edge, as is the case of exon 1 and 2. Exon 2 ends one base into the codon (in frame 1), which is drawn using an arrow on the end of the exon. Exon 3 starts in frame 1, and is drawn with a notch on the left side. This is the other way around for the boundary between exons 3 and 4, which is in frame 2.

Since the exons of a transcript should fit together, exons in conflicting frames (e.g., because of a frame shift inducing variant) are easily spotted because of their non-fitting boundaries.

The output of ExonViz is an SVG figure generated using the svg-py library, which can be used directly or modified using modern graphical editing programs. It is also possible to output the transcript and variants in TSV format, edit the transcript using any text editor or spreadsheet program, and draw the modified transcript using ExonViz. The online documentation has a number of examples of custom transcripts that can be visualized this way.

## Results

ExonViz is an application which allows users to automatically draw biologically accurate transcripts, including features like coding and non coding regions, exon reading frames and genetic variants. ExonViz can be employed to draw transcripts from any species of which the transcript information can be retrieved (Lefter et al., 2021), requiring only a gene, transcript or specific variant(s) as input.

More advanced illustrations such as the one in Figure 1C can be generated by using the command line version of ExonViz. This figure contains all ClinVar^7^ (Landrum et al., 2014) variants in the coding region of transcript NM_001378743.1 of *CYLD*, grouped into three categories (Landrum et al., 2024). Colors were chosen to be distinguishable by color blind people, according to Wong (2011). Exon 18 has been split over two rows, since it would not fit on a single row. This transcript has to be drawn at a scale of at least 1.2, since exon 6 is very small. Notice how exon 16 contains a large number of pathogenic variants, indicating a mutational hotspot. Finally, the figure illustrates how ExonViz collapses the legend if multiple variants have the same name and color.

Figure 1D contains a section of the Dp427m transcript of *DMD*, the largest human gene, containing 79 exons. The region shown covers exons 42 to 54, which can be targeted by various exon skipping therapies. Here, the scale has been reduced to 0.3 (each pixel represents three base pairs), to condense the figure while still showing the reading frames and the exon sizes to scale. In addition, the gap between the exons has been increased for clarity.

If the start frame is the same as the end frame for an exon (in other words, if the length of the exon is divisible by three), the exon may be skipped without disrupting the reading frame. This can be seen in Figure 1D for exons 47, 48 and 49. Note that although all in frame exons in Figure 1D are in frame 0, any exon with a coding length divisible by three is in frame.

Casimersen is an exon skipping therapy for Duchenne Muscular Dystrophy that skips exon 45 (Wagner et al., 2021) of the *DMD* transcript. As can be seen from Figure 1E, this would cause a frame shift in the wild type version of the transcript. But for patients with a deletion of exon 44, which in itself is a frame shift mutation, skipping exon 45 restores the reading frame, since exon 43 and 46 fit together. Likewise, Eteplirsen induces skipping of exon 51, restoring the reading frame for patients lacking exon 50 (Lim et al., 2017), as indicated in Figure 1F. Viltolarsen (Dhillon, 2020) and Golodirsen (Anwar & Yokota, 2020) skip exon 53, which can restore the reading frame for patients lacking exon 52. As can be seen from Figure 1G, in Viltolarsen and Golodirsen can also be used to treat deletions of 43-52, 45-52, 47-52, 48-52, 49-52 and 50-52 (Anwar & Yokota, 2020).

## Conclusion

To our knowledge, ExonViz is the first publicly accessible application that allows for automatic visualization of transcripts with additional features such as reading frames and variants along the transcript. ExonViz can be used for illustrations within publications, assessment of treatment options, for teaching purposes and genetic counseling. Figures generated by ExonViz are free to use under the Creative Commons BY license^4^. Furthermore, we allow the user to construct their own transcripts to incorporate features like poison or cryptic exons and alternative isoforms. ExonViz can be accessed as a web application via exonviz.rnatherapy.nl^1^ or installed via PyPI^2^. The source code is available on Github^5^.

## Data Availability

All data produced in the present study are available upon reasonable request to the authors

https://exonviz.rnatherapy.nl/

https://exonviz.readthedocs.io/en/latest/

https://github.com/DCRT-LUMC/exonviz

## Acknowledgments

We would like to thank the members of the Dutch Center for RNA Therapeutics for their ideas, suggestions and their feedback on earlier versions of ExonViz. We also thank Maximilian Haeussler and his colleagues at the UCSC for their efforts implementing exon frame information into the UCSC Genome Browser. We also thank Nanieke van den Berg for creating the figure for this manuscript.

## Footnotes

1 https://exonviz.rnatherapy.nl

2 https://pypi.org/project/exonviz

3 https://exonviz.readthedocs.io

4 https://creativecommons.org/licenses/by/4.0

5 https://github.com/DCRT-LUMC/exonviz

6 https://exonviz.readthedocs.io/en/latest/examples.html

7 https://www.ncbi.nlm.nih.gov/clinvar

## References

Alasoo, K. (2017). Wiggleplotr: Make read coverage plots from bigwig files. Bioconductor. doi, 10, B9.

Anwar, S., & Yokota, T. (2020). Golodirsen for duchenne muscular dystrophy. Drugs of Today (Barcelona, Spain: 1998), 56(8), 491–504. 10.1358/dot.2020.56.8.3159186

Cheerie, D., Meserve, M. M., Beijer, D., Kaiwar, C., Newton, L., Tavares, A. L. T., Verran, A. S., Sherrill, E., Leonard, S., Sanders, S. J., et al. (2025). Consensus guidelines for assessing eligibility of pathogenic dna variants for antisense oligonucleotide treatments. The American Journal of Human Genetics. 10.1016/j.ajhg.2025.02.017

Dhillon, S. (2020). Viltolarsen: First approval. Drugs, 80(10), 1027–1031. 10.1007/s40265-020-01339-3

Ferstay, J. A., Nielsen, C. B., & Munzner, T. (2013). Variant view: Visualizing sequence variants in their gene context. IEEE transactions on visualization and computer graphics, 19(12), 2546– 2555. 10.1109/TVCG.2013.214

Gustavsson, E. K., Zhang, D., Reynolds, R. H., Garcia-Ruiz, S., & Ryten, M. (2022). Ggtranscript: An r package for the visualization and interpretation of transcript isoforms using ggplot2. Bioinformatics, 38(15), 3844–3846. 10.1093/bioinformatics/btac409

Harrison, P. W., Amode, M. R., Austine-Orimoloye, O., Azov, A. G., Barba, M., Barnes, I., Becker, A., Bennett, R., Berry, A., Bhai, J., Bhurji, S. K., Boddu, S., BrancoÂ Lins, P. R., Brooks, L., Ramaraju, S. B., Campbell, L. I., Martinez, M. C., Charkhchi, M., Chougule, K., … Yates, A. D. (2023). Ensembl 2024. Nucleic Acids Research, 52(D1), D891–D899. 10.1093/nar/gkad1049

Landrum, M. J., Lee, J. M., Riley, G. R., Jang, W., Rubinstein, W. S., Church, D. M., & Maglott, D. R. (2014). Clinvar: Public archive of relationships among sequence variation and human phenotype. Nucleic acids research, 42(D1), D980–D985. 10.1093/nar/gkt1113

Landrum, M. J., Lee, J. M., Riley, G. R., Jang, W., Rubinstein, W. S., Church, D. M., & Maglott, D. R. (2024). Clinvar website. Retrieved June 17, 2024, from https://www.ncbi.nlm.nih.gov/clinvar

Lefter, M., Vis, J. K., Vermaat, M., den Dunnen, J. T., Taschner, P. E. M., & Laros, J. F. J. (2021). Mutalyzer 2: next generation HGVS nomenclature checker. Bioinformatics, 37 (18), 2811–2817. 10.1093/bioinformatics/btab051

Lim, K. R. Q., Maruyama, R., & Yokota, T. (2017). Eteplirsen in the treatment of duchenne muscular dystrophy. Drug design, development and therapy, 533–545. 10.2147/DDDT.S97635

Morales, J., Pujar, S., Loveland, J. E., Astashyn, A., Bennett, R., Berry, A., Cox, E., Davidson, C., Ermolaeva, O., Farrell, C. M., et al. (2022). A joint ncbi and embl-ebi transcript set for clinical genomics and research. Nature, 604(7905), 310–315. 10.1038/s41586-022-04558-8

Mühlhausen, S., Hellkamp, M., & Kollmar, M. (2015). Genepainter v. 2.0 resolves the taxonomic distribution of intron positions. Bioinformatics, 31(8), 1302–1304. 10.1093/bioinformatics/btu798

O’Leary, N. A., Wright, M. W., Brister, J. R., Ciufo, S., Haddad, D., McVeigh, R., Rajput, B., Robbertse, B., Smith-White, B., Ako-Adjei, D., et al. (2016). Reference sequence (refseq) database at ncbi: Current status, taxonomic expansion, and functional annotation. Nucleic acids research, 44(D1), D733–D745. 10.1093/nar/gkv1189

Pozo, F., Rodriguez, J. M., Vázquez, J., & Tress, M. L. (2022). Clinical variant interpretation and biologically relevant reference transcripts. NPJ Genomic Medicine, 7 (1), 59. 10.1038/s41525-022-00329-6

Richards, S., Aziz, N., Bale, S., Bick, D., Das, S., Gastier-Foster, J., Grody, W. W., Hegde, M., Lyon, E., Spector, E., et al. (2015). Standards and guidelines for the interpretation of sequence variants: A joint consensus recommendation of the american college of medical genetics and genomics and the association for molecular pathology. Genetics in medicine, 17 (5), 405–423. 10.1038/s41525-022-00329-6

Wagner, K. R., Kuntz, N. L., Koenig, E., East, L., Upadhyay, S., Han, B., & Shieh, P. B. (2021). Safety, tolerability, and pharmacokinetics of casimersen in patients with d uchenne muscular dystrophy amenable to exon 45 skipping: A randomized, double-blind, placebo-controlled, dose-titration trial. Muscle & nerve, 64(3), 285–292. 10.1038/gim.2015.30

Walker, L. C., de la Hoya, M., Wiggins, G. A., Lindy, A., Vincent, L. M., Parsons, M. T., Canson, D. M., Bis-Brewer, D., Cass, A., Tchourbanov, A., et al. (2023). Using the acmg/amp framework to capture evidence related to predicted and observed impact on splicing: Recommendations from the clingen svi splicing subgroup. The American Journal of Human Genetics, 110(7), 1046–1067. 10.1016/j.ajhg.2023.06.002

Wong, B. (2011). Points of view: Color blindness.

